# Solid fuel use status in the household and the risk of elevated blood pressure: findings from the 2017/18 Bangladesh Demographic and Health Survey

**DOI:** 10.1101/2022.06.04.22275991

**Authors:** Diba Paul, Dia Chowdhury, Hazrat Ali, Md. Syful Islam, Md Mostaured Ali Khan, Md. Nuruzzaman Khan

## Abstract

**Introduction:** Prevalence of hypertension is now increasing rapidly in Bangladesh, particularly among the socio-economically disadvantaged population. This could be linked to their higher use of solid fuel; however, related evidence is scarce in Bangladesh. We aimed to determine the associations of household solid fuel use and its exposure level with systolic blood pressure, diastolic blood pressure and hypertension.

**Methods:** Total of 7,320 women’s data extracted from the 2017/18 Bangladesh Demographic and Health Survey were analysed. We considered three outcome variables: (i) systolic blood pressure (continuous response), (ii) diastolic blood pressure (continuous response) and (iii) hypertension status (yes, no). Cooking fuel use (*clean fuel vs solid fuel*) and levels of exposure to household air pollution (HAP) through solid fuel use (unexposed, moderately exposed, highly exposed) were our primary exposure of interest. A multilevel mixed-effects Poisson regression model with robust variance was used to determine the association between exposure and outcome variable adjusting for confounders.

**Results:** Around 82% of the total respondents analysed used solid fuel for cooking. The overall age-standardised prevalence of hypertension was 28%. The likelihood of becoming hypertension was found 1.44 times (95% CI, 1.04-1.89) higher among respondents who used solid fuel as compared to the respondents who used clean fuel. The likelihood of hypertension was found to be increased with the increased exposure to HAP through the solid fuel used; 1.61 times (95% CI, 1.07-2.20) higher among the moderate exposure group and 1.80 times (95% CI, 1.27-2.32) higher among higher exposure group as compared to the women who used clean fuel. Similar associations were reported for systolic blood pressure and diastolic blood pressure.

**Conclusion:** Solid fuel use elevate systolic blood pressure, diastolic blood pressure and increases the likelihood of becoming hypertensive. Policies and programs are important to increase awareness about the adverse effects of solid fuel use on health, including hypertension. Focus should also be given to reducing solid fuel use and ensuring proper ventilation at the solid fuel use place.

## Background

Hypertension or elevated blood pressure (BP) (beyond 130/80 mmHg) is a serious global health concern with an estimated 1.28 billion people aged 30-79 years [1], a number that has been doubled in the last 30 years [2]. Two-thirds of them live in low- and middle-income countries (LMICs) [1, 2], mostly in South Asia and sub-Saharan Africa [2]. In 2019, the global prevalence of hypertension was 32% in women and 34% in men [2]. However, this prevalence is not the same across the life cycle and it is found that hypertension is less common in younger women as compared to their male counterparts, while hypertension is more common in elderly women than their men counterparts [3-5]. Hypertension is found to be associated with an increased prevalence of cardiovascular diseases, including stroke, myocardial infarction, and coronary heart disease [5, 6]. These are the major causes of mortality in LMICs including in Bangladesh. Importantly, over 90% of these deaths are premature and in number, this contributes to around 10.4 million deaths in a year [7]. Moreover, hypertension is found to be associated with 218 million disability-adjusted life years. It is therefore posing a country with a rising disease burden. Additionally, the current round of world development goals, the Sustainable Development Goals, targets to reduce one-third of the current occurrence of pre-mature deaths by 2030. However, it is unlikely that LMICs will achieve this target unless the current occurrence of pre-mature deaths due to rising blood pressure is controlled through proper management of hypertension.

The management of hypertension, which includes awareness, treatment and control of hypertension, is low worldwide, particularly in LMICs [5]. Globally, around 41% of the total hypertensive people do not know about their hypertensive status and the prevalence is higher in South Asia (55%) [2]. Even the respondents who know their hypertensive status, a higher percentage of them do not receive treatment and/or depend on ineffective or traditional medicines to control hypertension [8]. Therefore, a significant percentage of the diagnosed hypertension is uncontrolled. The causes are lower awareness of hypertension and its adverse effects. Moreover, controlling hypertriton needs continuous care which might be a burden for the many hypertensive people, particularly in low wealth quintile people, in LMICs given there is no universal healthcare coverage. Such lower management of hypertension is even higher among women because of lower knowledge about hypertension and poor access to healthcare services.

Globally, air pollution is a major determinant of hypertension [9, 10]. However, the proximity of adverse effects could vary depending upon the type of air pollution and exposure time [11, 12]. About 2.6 billion people worldwide use solid fuels for cooking, and a majority of them live in LMICs [13]. Such a higher prevalence emits considerably large amounts of health-damaging airborne pollutants, including particulate matter (PM), carbon monoxide, and polycyclic hydrocarbons [14]. As a result, world estimates show that 4 million people die each year due to HAP [13] and the relevant risk of dying has been found higher among people who use indoor solid fuel with poor ventilation [13].

In LMICs, women and children are highly exposed to HAP due to women’s role in household chores, cooking, and caring for infants [15]. They spend about four hours per day in the kitchen [16], with a possible root of long-time exposure because of no system of flues or hood to move out the smoke from the living place. This form of HAP is therefore found to be linked with the rising chorionic conditions, including hypertension, in LMICs and Bangladesh [17]. However, the existing studies have determined the association between solid fuel use or none use with hypertension, ignoring the place of using solid fuel instead of its importance. The reasons are indoor use of solid fuel can bring serve consequences, including developing hypertension, then outdoor use of solid fuel. This is found true in recent studies conducted in Myanmar and Bangladesh where likelihoods of under-five mortality were reported higher among the children whose mothers use solid fuel in an indoor place than solid fuel use in an outdoor place or none use of solid fuel [15]. In addition, related available studies in Bangladesh are used a less precious method to estimate the association between HAP and hypertension, such as the multivariate logistic regression model was used for clustering data with prevalence of hypertension >10, whereas the multilevel Poisson regression model is the most appropriate approach [17] and inadequate list of confounding factors. Therefore, true understating of the strengths of the associations between HAP and hypertension are still lacking. We, therefore, conducted this study to determine the associations of exposure to HAP and the level of exposure to HAP with hypertension in Bangladesh using nationally representative survey data.

## Methods

### Study design

Data for this study was extracted from the 2017/18 Bangladesh Demographic and Health Survey. This is the most recent survey in Bangladesh conducted as part of the Demography and Health Survey Program of the USA. The Ministry of Health and Family Welfare of Bangladesh conducted this survey through its affiliated institutes, the National Institute of Population Research and Training. International development partners, including UNICEF and UNDP, have supported this research financially and technically. The survey followed a two-stage stratified random sample technique to collect a nationally-representative sample. In the first phase, 675 primary sampling units (PSUs) were randomly selected from 293,579 PSUs generated by the Bangladesh Bureau of Statistics as part of conducting the 2011 National Population Census. Data was collected from the 672 PSUs of them. A fixed number of 30 households were selected from each PSU in the second stage of sampling. This produced a list of 20,160 households for data collection, while data were collected from 19,457 households with a 96.5% inclusion rate. Further sampling was done to collect blood pressure (BP) measurements whereas one-fourth of the selected households (7 to 8 households per PSU) were chosen, which produced 4,864 households. There were 14,704 respondents aged 18 years or more in these selected households; 12,924 of them had BP measured with a response rate of 87.9%. Details of the sampling procedure have been published elsewhere [18].

### Study outcomes

This study was conducted for three outcomes: (i) systolic blood pressure (continuous variable, reported in millimetres of mercury [mmHg]), (ii) diastolic blood pressure (continuous variable, reported in millimetres of mercury [mmHg]), and (iii) hypertension status (yes, no). The survey reported systolic blood pressure (SBP) and diastolic blood pressure (DBP) of the included respondents during the survey. For this, the survey used digital oscillometer BP measuring devices with automatic upper-arm inflation and automatic pressure releases devices [18]. BP was checked three times with an interval of at least 5 minutes, and the average values of the second and third measurements were used as the individual BP [18]. We classified an individual as hypertensive if he/she had systolic blood pressure ≥140 mmHg and/or a diastolic blood pressure (DBP) ≥90 mmHg or reported he/she currently using any antihypertensive drugs to control BP. This is the recommendation of the National Guidelines for Management of Hypertension in Bangladesh and is comparable with the 2018 European Society of Hypertension and European Society of Cardiology hypertension guidelines [19].

### Exposure variables

Two exposure variables were considered: (i) type of cooking fuel used (solid fuel, clean fuel) and (ii) level of exposure to HAP through solid fuel used (unexposed, moderately exposed, highly exposed). The BDHS recorded the type of fuel used in the respondents’ households for cooking by asking “*What type of fuel does your households mainly used for cooking?*”. A list of fuels was provided to give the response. In case, the fuel used by the respondents was not on the list, they were allowed to write the name of the fuel. We reclassified these responses as solid fuel used (if the respondents recorded coal, lignite, charcoal, wood, straw, shrubs, grass, agricultural crop and animal dung) and clean fuel used (electricity, liquid petroleum gas, natural gas and biogas) to generate the first exposure variable. Respondents were also asked about the place of cooking in their households by asking “*Is the cooking usually done in the house, in a separate building, or outdoors?*”. Responses recorded for this question were considered along with the type of cooking fuel respondents used to generate the second exposure variable. The respondents were considered *unexposed* if respondents recorded clean fuels use in their households for cooking purposes; *moderately exposed*, if respondents recorded solid fuels use in their households for cooking purpose, however, the cooking conducts in a separate building or outdoors; and *highly exposed*, if respondents recorded solid fuels use in their households for cooking purpose, however, the cooking conducts inside their houses. We generate this variable by following previous studies conducted in LMICs [8, 17-19, 21, 26-28].

### Confounding adjustment

Individual-, household-, and community-level factors were considered confounding factors. We first generate a list of confounding variables by reviewing the relevant studies conducted in LMICs, particularly in Bangladesh [8, 17-20, 22, 23, 29]. The availability of the data of the selected variables in the survey we analysed was then checked. The variables whose data were found to be available in the survey were then analysed with the forward regression analysis. The variables that were found significant, as presented in Table 3, were considered as confounding factors.

### Statistical analysis

Descriptive statistics was used to describe the characteristics of the respondents. A separate multilevel mixed-effects Poisson regression model was used to assess the associations of type of cooking fuels use and exposure level of HAP through using solid fuel with systolic blood pressure, diastolic blood pressure and hypertensive status. The reason for using this model was the nesting structure of the BDHS data and the higher prevalence (>10%) of hypertension. Previous studies found in such cases multilevel Poisson regression model produces more precise results [30]. Survey weight was also considered in the model. We ran both unadjusted and adjusted models. In the unadjusted model, only the particular exposure and outcome variable was considered. The confounding variables considered were added with the particular exposure and outcome variable in the adjusted model. Results were recorded as Prevalence Ratio (PR) with its 95% Confidence Interval (95% CI). We performed all descriptive statistics using the Stata software version 15.1 (Stata corporation, college station, Texas, USA).

## Results

Background characteristics of the respondents are presented in Table 1. The mean age of the respondents was 38 years and the mean year of education was 4.87 years. Nearly 82% of the total respondents were found to be used solid fuel for household cooking. The major form of solid fuels was wood (46.82%) following agricultural crops (27.21%) and animal manure (7.12%). Around three-fourths of the total respondents reported they used a separate building for cooking. Only 2.10% of the total respondents reported they used an indoor place for cooking. Once we considered cooking fuel and place together to create an index of level of exposure to HAP through cooking fuels, we found around 81% of the total respondents were moderately exposed to HAP and only 0.58% of the total respondents were highly exposed to HAP. The mean systolic blood pressure was 121 and the mean diastolic blood pressure was 80.26. The prevalence of hypertension was 28.25%.

**Table 1:**
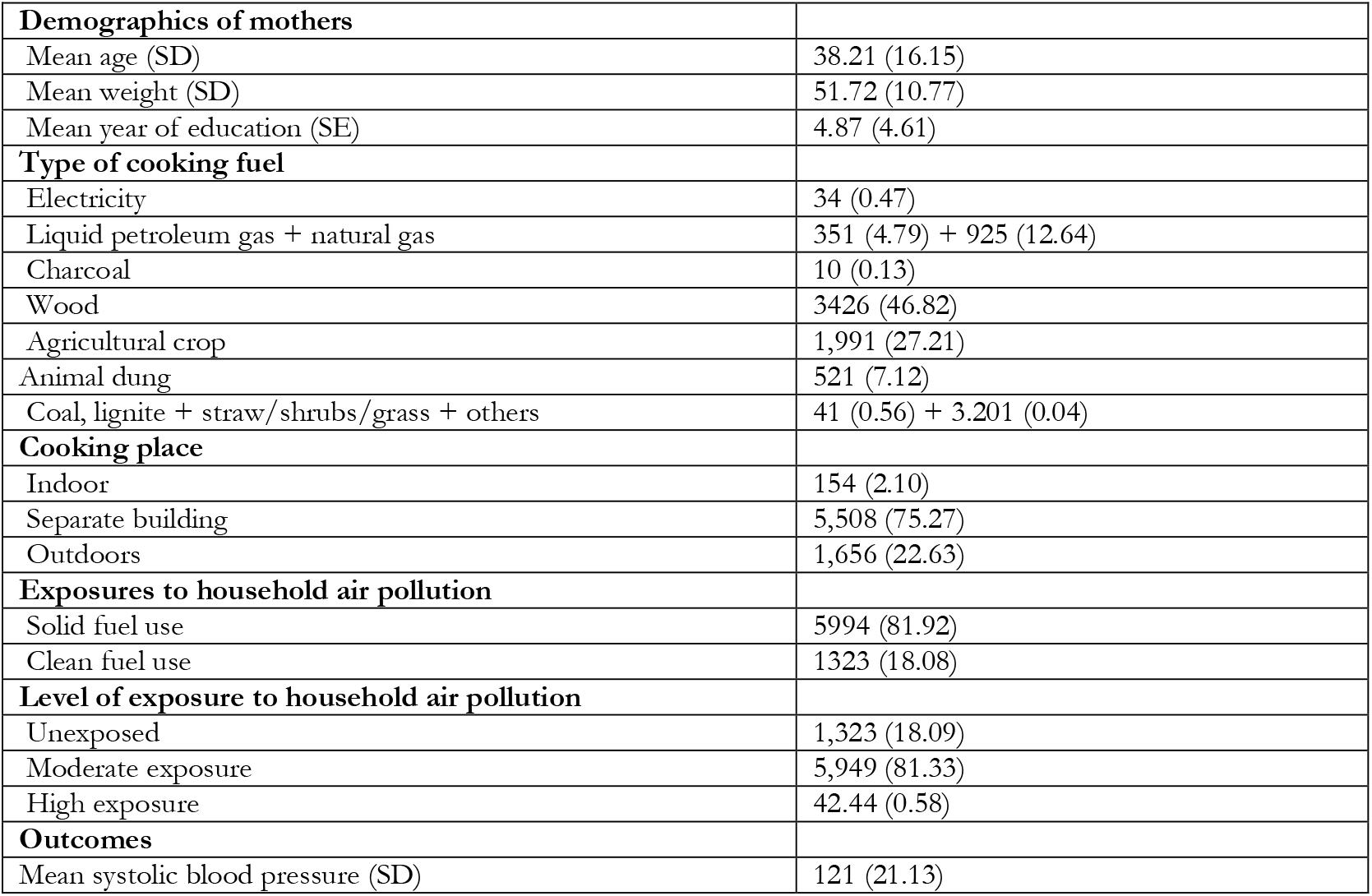
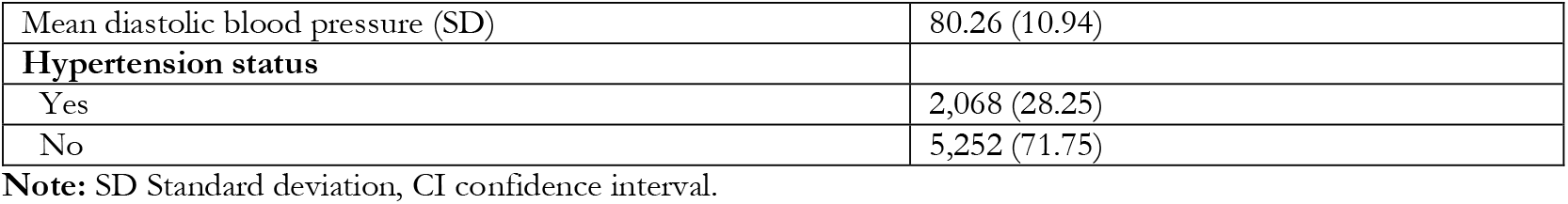
Key information about the study participants, exposure, and outcome variables.

In **Table 2**, the crude and age-standardized prevalence of **hypertension** are presented by respondents’ socio-demographic characteristics. The age-adjusted prevalence of hypertension was relatively high among illiterate women (44%) than higher educated women (16%). Prevalence of hypertension was found to be increased with the increase in respondents’ age and more than half of the respondents aged 50 years and more were found hypertensive. Prevalence of hypertension was also found to be increased with the increase in body mass index, from a 20% prevalence among underweighted women to 44% among obese women. At the divisional level, women who resided in the Rangpur and Chattogram reported a higher prevalence of hypertension.

**Table 2:**
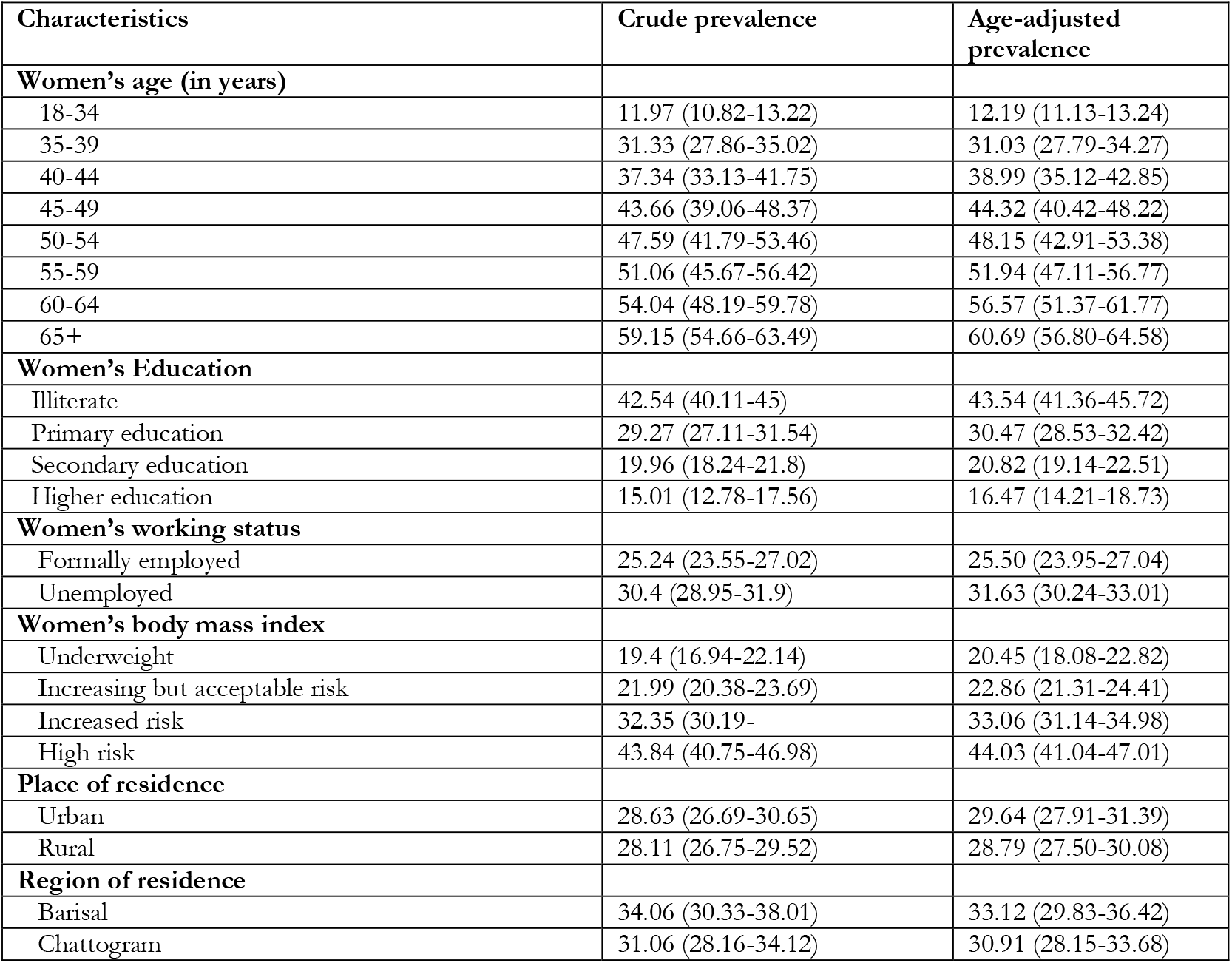
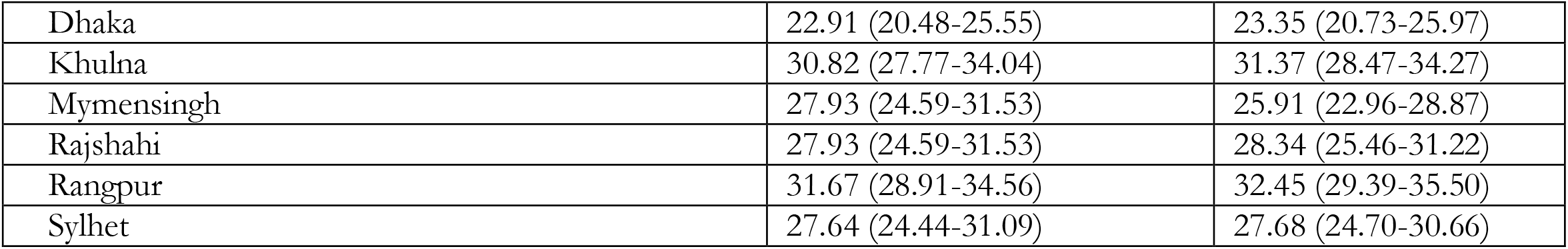
Crude and age-standardized prevalence of hypertension by respondents’ socio-demographic characteristics, BDHS, 2017/18.

| Characteristics | Crude prevalence | Age-adjusted prevalence |
| --- | --- | --- |
| **Women's age (in years)** |  |  |
| 18-34 | 11.97 (10.82-13.22) | 12.19 (11.13-13.24) |
| 35-39 | 31.33 (27.86-35.02) | 31.03 (27.79-34.27) |
| 40-44 | 37.34 (33.13-41.75) | 38.99 (35.12-42.85) |
| 45-49 | 43.66 (39.06-48.37) | 44.32 (40.42-48.22) |
| 50-54 | 47.59 (41.79-53.46) | 48.15 (42.91-53.38) |
| 55-59 | 51.06 (45.67-56.42) | 51.94 (47.11-56.77) |
| 60-64 | 54.04 (48.19-59.78) | 56.57 (51.37-61.77) |
| 65+ | 59.15 (54.66-63.49) | 60.69 (56.80-64.58) |
| **Women's Education** |  |  |
| Illiterate | 42.54 (40.11-45) | 43.54 (41.36-45.72) |
| Primary education | 29.27 (27.11-31.54) | 30.47 (28.53-32.42) |
| Secondary education | 19.96 (18.24-21.8) | 20.82 (19.14-22.51) |
| Higher education | 15.01 (12.78-17.56) | 16.47 (14.21-18.73) |
| **Women's working status** |  |  |
| Formally employed | 25.24 (23.55-27.02) | 25.50 (23.95-27.04) |
| Unemployed | 30.4 (28.95-31.9) | 31.63 (30.24-33.01) |
| **Women's body mass index** |  |  |
| Underweight | 19.4 (16.94-22.14) | 20.45 (18.08-22.82) |
| Increasing but acceptable risk | 21.99 (20.38-23.69) | 22.86 (21.31-24.41) |
| Increased risk | 32.35 (30.19- | 33.06 (31.14-34.98) |
| High risk | 43.84 (40.75-46.98) | 44.03 (41.04-47.01) |
| **Place of residence** |  |  |
| Urban | 28.63 (26.69-30.65) | 29.64 (27.91-31.39) |
| Rural | 28.11 (26.75-29.52) | 28.79 (27.50-30.08) |
| **Region of residence** |  |  |
| Barisal | 34.06 (30.33-38.01) | 33.12 (29.83-36.42) |
| Chattogram | 31.06 (28.16-34.12) | 30.91 (28.15-33.68) |
| Dhaka | 22.91 (20.48-25.55) | 23.35 (20.73-25.97) |
| Khulna | 30.82 (27.77-34.04) | 31.37 (28.47-34.27) |
| Mymensingh | 27.93 (24.59-31.53) | 25.91 (22.96-28.87) |
| Rajshahi | 27.93 (24.59-31.53) | 28.34 (25.46-31.22) |
| Rangpur | 31.67 (28.91-34.56) | 32.45 (29.39-35.50) |
| Sylhet | 27.64 (24.44-31.09) | 27.68 (24.70-30.66) |

Table 3 shows the unadjusted and adjusted associations of household air pollution with systolic blood pressure, diastolic blood, and hypertension status. The likelihoods of systolic blood pressure (aPR, 1.42, 95% CI 1.01-1.84), diastolic blood pressure (aPR, 1.76, 95% CI 1.04-2.54) and hypertension (aPR, 1.44, 95% CI 1.04-1.89) were found higher among women who used solid fuel as compared to their counterparts who used clean fuel for cooking. The likelihoods were even found higher once we considered the augmented measure of exposure to HAP. Compared to the unexposed women, a significant increase in the likelihood of systolic blood pressure, diastolic blood pressure and hypertension was reported among women moderately exposed to the HAP and highly exposed to the HAP. We found likelihoods of systolic blood pressure (aPR, 1.36, 95% CI 1.03-2.06), diastolic blood pressure (aPR, 1.86, 95% CI 1.04-2.70) and hypertension (aPR, 1.61, 95% CI 1.07-2.20) were 1.36 to 1.86 times higher among women moderately exposed to HAP than unexposed to HAP. This was found further higher among highly exposed women as compared to the unexposed with likelihoods of 2.56 (95% CI 1.40-3.70) times higher for systolic blood pressure, 2.49 (95% CI, 1.20-3.78) times higher for diastolic blood pressure and 1.80 (95% CI, 1.27-2.32) times higher for hypertension.

**Table 3:** Unadjusted **and adjusted** association of household air pollution with systolic blood pressure, diastolic blood, and hypertension status, Bangladesh

|  | Systolic blood pressure,<br>PR, 95% CI |  | Diastolic blood pressure<br>PR, 95% CI |  | Hypertension<br>PR, 95% CI |  |
| --- | --- | --- | --- | --- | --- | --- |
|  | Unadjusted | Adjusted | Unadjusted | Adjusted | Unadjusted | Adjusted |
| **Cooking fuel** |  |  |  |  |  |  |
| Clean fuel (ref) | 1.00 | 1.00 | 1.00 | 1.00 | 1.00 | 1.00 |
| Solid fuel | 1.52 (1.00-2.03) | 1.42 (1.01-1.84) | 1.84 (1.06-2.54) | 1.76 (1.04-2.54) | 1.59 (1.05-2.14) | 1.44 (1.04-1.89) |
| **Level of exposure to household air pollution** |  |  |  |  |  |  |
| Unexposed | 1.00 | 1.00 | 1.00 | 1.00 | 1.00 | 1.00 |
| Moderate exposure | 1.42 (1.06-1.84) | 1.36 (1.03-2.06) | 2.11 (1.09-3.12) | 1.86 (1.04-2.70) | 1.79 (1.09-2.14) | 1.61 (1.07-2.20) |
| High exposure | 2.63 (1.48-3.78) | 2.56 (1.40-3.70) | 2.78 (1.28-4.17) | 2.49 (1.20-3.78) | 2.70 (1.11-4.08) | 1.80 (1.27-2.32) |

## Discussion

In this study, we explored the current use of solid fuel use in Bangladesh and associations of solid fuel used with systolic blood pressure, diastolic blood pressure and hypertension. For this, along with the household solid fuel using status, we developed an augmented index, exposure level to HAP, by combining places where solid fuel are used and determined their associations with the systolic blood pressure, diastolic blood pressure and hypertension status by using the multilevel Poisson regression model adjusting for possible confounders. We found solid fuel use increases the risk of becoming hypertensive as well as increases the systolic and diastolic blood pressure. The likelihood was even found higher among women who used solid fuel in the indoor place than the outdoor place or use clean fuel. These findings give an important pathway of the rising prevalence of hypertension in Bangladesh with its current prevalence of 80% for solid fuel use, 2% of them used solid fuel in an indoor place. Besides, this study’s findings are robust in response to the study data analysed, the type of confounding factors considered, and the statistical methods used. Therefore, the findings are expected to help policymakers to create evidence-based policies and programs in Bangladesh that will accelerate its progress in line with achieving the SDGs by 2030.

The direction of the associations of solid fuel used and exposure level to HAP through solid fuel used with systolic blood pressure, diastolic blood pressure and hypertension that were reported in this study are consistent with previous studies conducted in LMICs including Bangladesh [17, 20-22]. This association arises because women in LMICs traditionally perform cooking activities and spend a significant time in the kitchen [22]. Consequently, they are exposed to black carbon along with a great number of air pollutants which are emitted by the solid fuel. These pollutants cause an acute increase in systolic and diastolic blood pressure among women, as well as make them hypertensive [23].

One important aspect of this study’s findings is the gradual increasing likelihood of systolic blood pressure, diastolic blood pressure and hypertension with the increased exposure level of HAP. As far as we know, this observation is very first in Bangladesh as well as LMICs, therefore, we could not compare our findings. However, recent studies in Bangladesh and Myanmar found an increased likelihood of under-five mortality with the increased exposure level of HAP [15, 24]. Increased exposure level to HAP means women used solid fuel inside their households and or living rooms mostly without a proper smoking ventilation system. Consequently, this increases their exposure time, even following the end of cooking. Moreover, traditionally Bangladeshi people cook three times a day and every time cooking takes around 2/3 hours. With the poor ventilation system, such usual 6/9 hours of direct exposure to HAP increases to 12/16 hours, considering after cooking hours HAP. This higher time exposure and breathing smoke could be associated with rising likelihoods of systolic blood pressure, diastolic blood pressure and hypertension with the exposure level to HAP, from unexposed to moderately or highly exposed.

Besides these, uneducated and rural women in Bangladesh, like other LMICs, mostly perform cooking in an indoor place. They usually do not know about the risk of using solid fuel. They also do not aware of the blood pressure and the importance of its management. As such, they usually do not check their blood pressure until they get sick. Such lack of awareness along with the rising level of exposure to HAP could be another important direction of the rising likelihood of systolic blood pressure, diastolic blood pressure and hypertension with the rising level of HAP exposure [23, 25-27].

The study has several strengths and some limitations. We conducted this study by analysing a large scale nationally representative survey data with advanced statistical modelling and adjusting for all possible confounders. Our multilevel mixed-effects Poisson regression model also addressed the issues of overestimation and/or overestimation that were produced in the conventional logistic regression model. Therefore, the findings of this study are robust and should be used in national-level policy and program making. However, as we analysed cross-sectional survey data, therefore, the relationships reported were correlational only, not casual. Moreover, all data in the survey, including the type of cooking fuels respondents used and the place where respondents used solid fuel, were self-reported, and interviewers couldn’t validate the responses. This could be an important source of bias response; however, any bias could be random. Besides, systolic blood pressure, diastolic blood pressure and hypertension are linked with many other factors, including food habits and behavioural factors. Therefore, their adjustment in the model is important to get accurate results. However, we could not do that because of the lack of data in the survey we analysed.

## Conclusion

We found HAP due to solid fuel use and increase exposure to HAP through solid fuel use increase the likelihood of systolic blood pressure, diastolic blood pressure, and hypertension. This indicates a challenge for Bangladesh considering 80% of the total Bangladeshi households use solid fuel for cooking and rising blood pressure is one of the major disease burdens in Bangladesh. Improved cooking facilities, ventilation systems and awareness about the adverse effects of solid fuel use are important to reduce the burden of hypertension in Bangladesh, particularly among women.

## Data Availability

The Demography and Health Survey (DHS) program of the USA is the custodian of 2017 BDHS data. It is freely available for the user upon submission of reasonable requests to the DHS.

https://dhsprogram.com/data/available-datasets.cfm

## Declaration

This study has not been published or presented anywhere before.

I confirm that the manuscript has been submitted solely to this journal and is not published, in-press, or submitted elsewhere.

I can confirm that all the research meets the ethical guidelines, including adherence to the legal requirements of the study country.

## Ethics

This study analysed secondary data publicly available. Ethical approval for this survey was provided by the Bangladesh Medical research council and the Demographic and Health Survey Program of the USA. No additional ethical approval is required to conduct this study.

## Consent for publication

Not applicable

## Authors’ contribution

Khan MN, Islam MS, and Khan MM designed this study. Paul D and Chowdhury D, and Ali H analyzed the data along. Paul D and Chowdhury D write the first draft of this manuscript. Khan MN, Islam MS, and Khan MM critically revised this manuscript. All authors approved this submitted version of the manuscript.

## Acknowledgement

We acknowledge the DHS program of the USA, custodian of the data used in this study, for approving to use of their data.

## Reference

1. WHO. More than 700 million people with untreated hypertension. 2021 [cited 2022 22 May]; Available from: https://www.who.int/news/item/25-08-2021-more-than-700-million-people-with-untreated-hypertension.

2. Zhou, B., et al., Worldwide trends in hypertension prevalence and progress in treatment and control from 1990 to 2019: a pooled analysis of 1201 population-representative studies with 104 million participants. The Lancet, 2021. 398(10304): p. 957–980.

3. Ahmad, A. and S. Oparil, Hypertension in women: recent advances and lingering questions. Hypertension, 2017. 70(1): p. 19–26.

4. Peltzer, K. and S. Pengpid, The prevalence and social determinants of hypertension among adults in Indonesia: a cross-sectional population-based national survey. International journal of hypertension, 2018. 2018.

5. Ahmad, A. and S. Oparil, Hypertension in Women. Hypertension, 2017. 70(1): p. 19–26.

6. Campbell, N.R., et al., Using the Global Burden of Disease study to assist development of nation-specific fact sheets to promote prevention and control of hypertension and reduction in dietary salt: a resource from the World Hypertension League. J Clin Hypertens (Greenwich), 2015. 17(3): p. 165–7.

7. Bennett, J.E., et al., NCD Countdown 2030: pathways to achieving Sustainable Development Goal target 3.4. The Lancet, 2020. 396(10255): p. 918–934.

8. Santosa, A., et al., Gender differences and determinants of prevalence, awareness, treatment and control of hypertension among adults in China and Sweden. BMC Public Health, 2020. 20(1): p. 1763.

9. Rafaj, P., et al., Outlook for clean air in the context of sustainable development goals. 2018. 53: p. 1–11.

10. Abba, M.S., et al., Household Air Pollution and High Blood Pressure: A Secondary Analysis of the 2016 Albania Demographic Health and Survey Dataset. International Journal of Environmental Research and Public Health, 2022. 19(5): p. 2611.

11. Chen, H., et al., Spatial association between ambient fine particulate matter and incident hypertension. Circulation, 2014. 129(5): p. 562–569.

12. Hamanaka, R.B. and G.M. Mutlu, Particulate matter air pollution: effects on the cardiovascular system. Frontiers in endocrinology, 2018. 9: p. 680.

13. WHO. Household air pollution and health. 2021 [cited 2022 12th May]; Available from: https://www.who.int/news-room/fact-sheets/detail/household-air-pollution-and-health.

14. Amegah, A.K., R. Quansah, and J.J. Jaakkola, Household air pollution from solid fuel use and risk of adverse pregnancy outcomes: a systematic review and meta-analysis of the empirical evidence. PLoS One, 2014. 9(12): p. e113920.

15. Rana, J., et al., Association between household air pollution and child mortality in Myanmar using a multilevel mixed-effects Poisson regression with robust variance. Scientific Reports, 2021. 11(1): p. 12983.

16. Jain, M. Addressing complexities of measuring women’s time use in Bangladesh. 2015 [cited 2022 14 May]; Available from: https://a4nh.cgiar.org/2015/02/02/addressing-complexities-of-measuring-womens-time-use-in-bangladesh/.

17. Khan, J.R., M.B. Hossain, and R.D. Gupta, Household cooking fuels associated with elevated blood pressure among adult women: a national-wide assessment in Bangladesh. Environmental Science and Pollution Research, 2021. 28(47): p. 67814–67821.

18. NIPORT, Bangladesh Demographic and Health Survey 2017-18. 2020, National Institute of Population, Research adn Training (NIPORT), Ministry of Health and Family Welfare: Dhaka, Bangladesh.

19. Chowdhury, M.Z.I., et al., Hypertension prevalence and its trend in Bangladesh: evidence from a systematic review and meta-analysis. Clinical Hypertension, 2020. 26(1): p. 10.

20. Khan, M.N., et al., Household air pollution from cooking and risk of adverse health and birth outcomes in Bangladesh: a nationwide population-based study. Environmental Health, 2017. 16(1): p. 1–8.

21. Li, L., et al., Indoor air pollution from solid fuels and hypertension: A systematic review and meta-analysis. Environmental Pollution, 2020. 259: p. 113914.

22. Bloomfield, G.S., et al., Waiting to inhale: An exploratory review of conditions that may predispose to pulmonary hypertension and right heart failure in persons exposed to household air pollution in low-and middle-income countries. Global heart, 2012. 7(3): p. 249–259.

23. Norris, C., et al., A panel study of the acute effects of personal exposure to household air pollution on ambulatory blood pressure in rural Indian women. Environmental Research, 2016. 147: p. 331–342.

24. Alam, M.B., et al., Household Air Pollution from Cooking Fuels and its Association with Under-Five Mortality in Bangladesh. medRxiv, 2022.

25. Young, B.N., et al., Exposure to household air pollution from biomass cookstoves and blood pressure among women in rural Honduras: A cross-sectional study. Indoor Air, 2019. 29(1): p. 130–142.

26. Baumgartner, J., et al., Indoor Air Pollution and Blood Pressure in Adult Women Living in Rural China. Environmental Health Perspectives, 2011. 119(10): p. 1390–1395.

27. Rajkumar, S., et al., Household air pollution from biomass-burning cookstoves and metabolic syndrome, blood lipid concentrations, and waist circumference in Honduran women: A cross-sectional study. Environmental Research, 2019. 170: p. 46–55.

